# Management strategies and prediction of COVID-19 by a fractional order generalized SEIR model

**DOI:** 10.1101/2020.06.18.20134916

**Authors:** Lihong Guo, Yanting Zhao, YangQuan Chen

**Affiliations:** Institute of Mathematics, Jilin University, Changchun 130012, P. R. China; Lab of Vibration Control & Vehicle Control Department of Automation, University of Science and Technology of China, Hefei 230022, P. R. China; Mechatronics, Embedded Systems and Automation Laboratory, Dept. of Engineering University of California, Merced, CA 95343, USA

**Keywords:** Fractional order generalized SEIR epidemic model, Prediction, Management strategies

## Abstract

In this project, we study a class of fractional order generalized SEIR epidemic models. Based on the public data from Jan. 22th to May 15th, 2020, we reliably estimate key epidemic parameters and make predictions on the peak point and possible ending time for the target region. We analyze the current management strategy and predict the future implementation of different management strategies. Numerical simulations which support our analysis are also given.

## 1 Introduction

Since the identification of coronavirus disease (COVID-19) in Wuhan, China in December 2019, the virus has spread rapidly around the world, resulting in a global pandemic. As of May 16th 21:30 PT 4,634,132 people in over 180 countries and regions have been infected [1]. As there is no drug treatment or vaccine, and it has a high infection rate, so there is no trend to slow down. According to the data on the Johns Hopkins web site: https://coronavirus.jhu.edu/data/mortality, the fatality rate in the United States is now 6.1%. Therefore, governments around the world are taking necessary measures to stop the epidemic of diseases, such as regional blockades, maintaining social distance and working from home.

Therefore, it is necessary to model the spreading trend of epidemic diseases and predict the future development trend. In this project, we study a class of fractional order generalized SEIR epidemic models with age groups. Based on the public data on the web site: https://midasnetwork.us/mmods/ from Jan. 22th to May 15th, 2020, we reliably estimate key epidemic parameters and make predictions on the peak point and possible ending time for the target region. We analyze the current management strategy and predict the future implementation of different management strategies. Numerical simulations which support our analysis are also given.

## 2 Model

Based on current local policies and conditions, we make the following assumptions:

During the COVID-19 outbreak, a US county of 100,000 people that pre-emptively initiated, and adhered to, stringent social distancing guidelines (i.e., full lockdown with workplace and school closures) until May 15th, 2020. In the meantime, current (i.e., partial) travel restrictions remain in place, so that no international importation is allowed and domestic importations are limited. In addition, we assume no local testing/contact tracing and isolation of infected individuals.

Currently, one of the most traditional and relatively simple mathematical frameworks for studying epidemics is the SEIR compartmental model. The model consists of four parts: S represents the number of susceptible cases, E represents the exposed cases, which means the number of individuals in the incubation period, I represents the number of infectious cases, and R represents the number of recovered or dead individuals.

In this paper, based on the SEIR model, a class of SEIR models with power law infection rate is considered, which is called fractional order generalized SEIR model. Generally, the infection rate of an infectious disease is assumed to be a constant. In paper [2], Singer studied the infection rate model of COVID-19 this time, showing that the infection rate follows the power law:

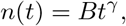

where *n*(*t*) is the number of infections at time *t, B* is a constant number, *γ* is the exponent. This research is also consistent with the phenomenon of human interactions and structures conforming to the small-world network phenomena and scale-free networks.

It can also be seen from the autocorrelation function of the number of confirmed cases that the autocorrelation function has the property of long memory, so the fractional order model can be used to describe the infection rate, see Fig. 2.1.

**Figure 2.1:**
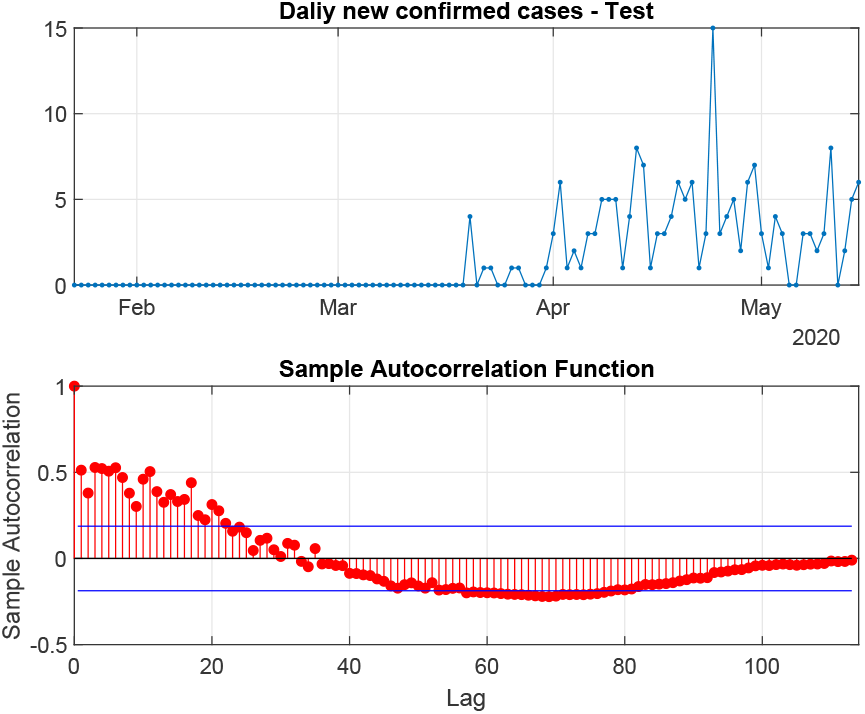
The autocorrelation function of daily new confirmed cases

**Figure 2.2:**
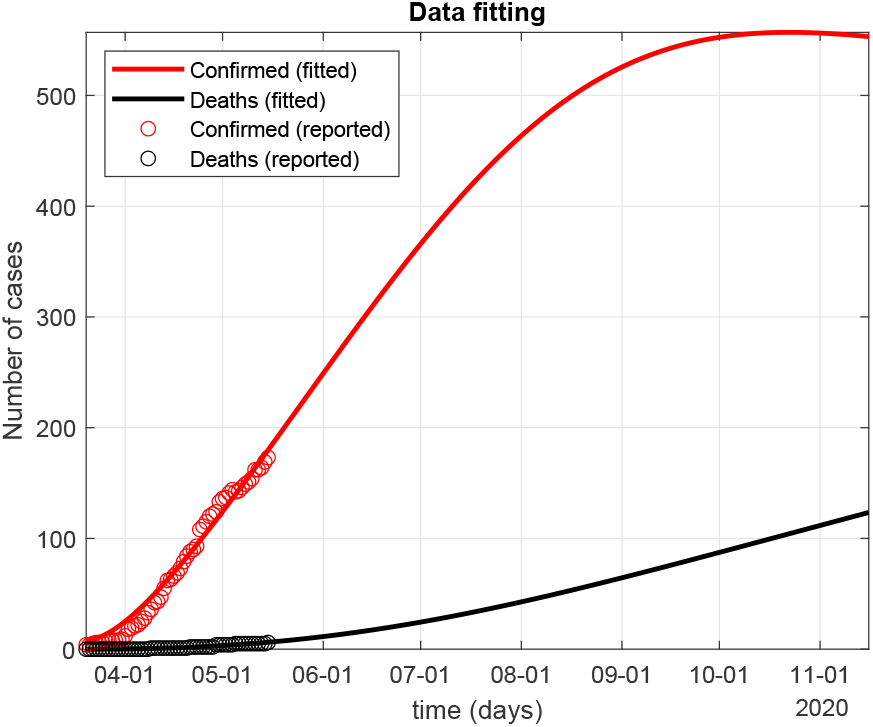
The fitting curve

### Definition 2.1.

[3] *The fractional integral of order α >* 0 *for a function f* (*t*) *is defined by*

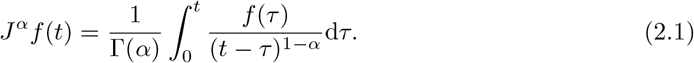

Based on the data currently available, we have added research on quarantined cases Q and death cases D based on the SEIR model with power law infection rate, named fractional order generalized SEIR model

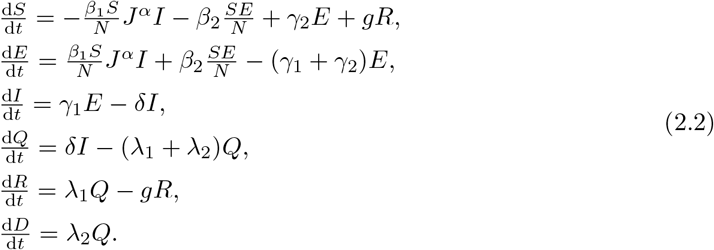

For short-term outbreaks of infectious diseases, the impact of population migration is generally not considered. The biological meanings of state variables and parameters are shown in Table 1 and Table 2. The range of values of some parameters in the model can be obtained from the existing literature [4], also shown in Table 2. For some parameter values that are difficult to measure, such as exposure rate *β*1 and *β*_2_, we obtain by fitting the given data. As shown in the Fig. 2.2, the data fitting shows that the peak of the current number of confirmed cases occurred around October 22, with about 557 people. The number of deaths has been increasing.

**Table 1:** The biological meanings of state variables for system (2.2)

| Variables | Description |
| --- | --- |
| $S$ | Number of susceptible individuals at time $t$ |
| $E$ | Number of exposed individuals at time $t$ |
| $I$ | Number of infectious individuals at time $t$ |
| $Q$ | Number of quarantined individuals at time $t$ |
| $R$ | Number of recovered individuals at time $t$ |
| $D$ | Number of death individuals at time $t$ |

**Table 2:**
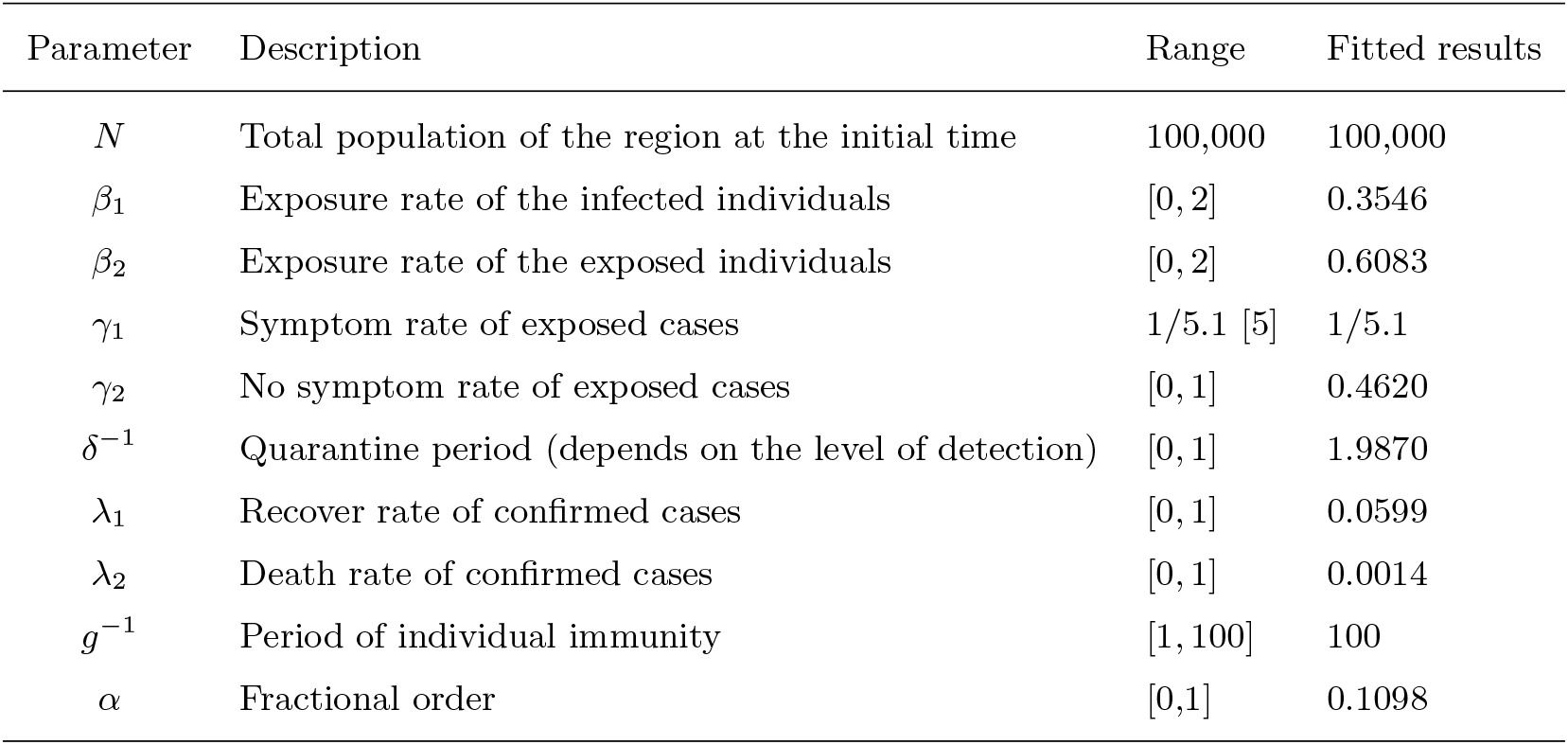
The biological meanings and values of parameters for system (2.2)

### Remark 2.1.

*The data fitting part is based on Simulink module. Based on the real data, we also use the Simulink Design Optimization toolbox to optimize the parameters in the equation*.

## 3 Interventions and predictions

In the current situation, policymakers, such as county chief executives, may be most concerned about formulating policies related to isolation and reopening in the coming months before the flu season begins [6]. These policies may directly affect public health, hospital resources and the local economy. Therefore, we will consider workplace-related relaxation and assume the following four intervention options

1. Continue with current workplace and school closures until November 15 (baseline full control scenario);
2. Relax current social distancing 2 weeks after peak: open workplaces only (schools remain closed through November 15);
3. Relax social distancing when the number of new daily cases is at 1% of peak: open workplaces only (schools remain closed through November 15);
4. Immediately relax all current restrictions on workplaces (schools remain closed through November 15).

To reflect the effects of these interventions, we use the following five outcomes (metrics) as an illustration

1. Cumulative number of infected individuals through November 15;
2. Cumulative number of deaths through November 15;
3. Peak hospitalizations through November 15;
4. Probability of a new local outbreak (more than 10 new cases/day) before November 15;
5. Total number of days workplaces closed through November 15.

Based on different intervention scenarios, the results are shown in Fig. 3.5, 3.7 and Table 3

**Table 3:** Projection outcomes/objectives with different interventions

|  | Closed | 2 weeks | 1 percent | Open |
| --- | --- | --- | --- | --- |
| cumu_infections | 3,231 | 21,585 | 3,231 | 64,648 |
| cumu_deaths | 123 | 1,132 | 123 | 3,300 |
| peak_hosp | 111 | 5,820 | 111 | 8,830 |
| prob_outbreak | 75.52% | 44.4% | 75.52% | 35.27% |
| days_closed | 184 | 170 | 184 | 0 |

**Figure 3.3:**
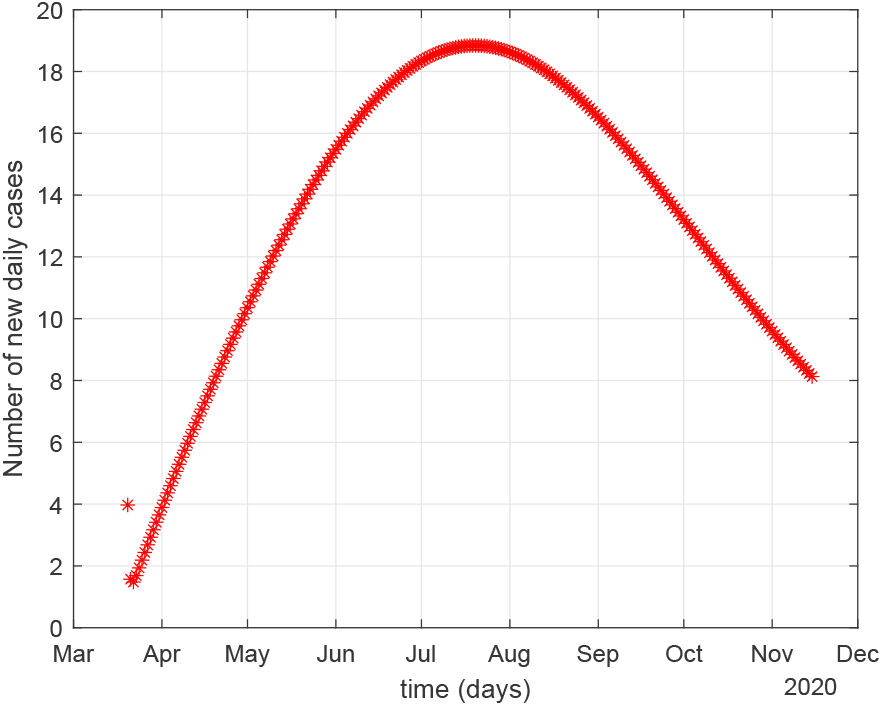
The number of new daily cases continue with current restrictions

**Figure 3.4:**
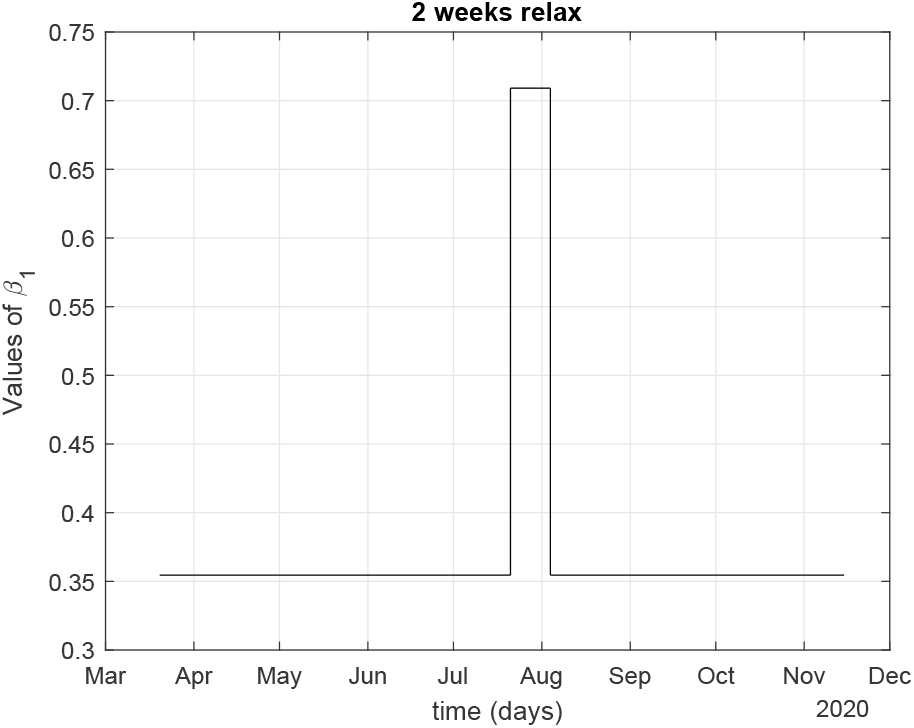
The values of *β*_1_ before and after 2 weeks of relaxation

**Figure 3.5:**
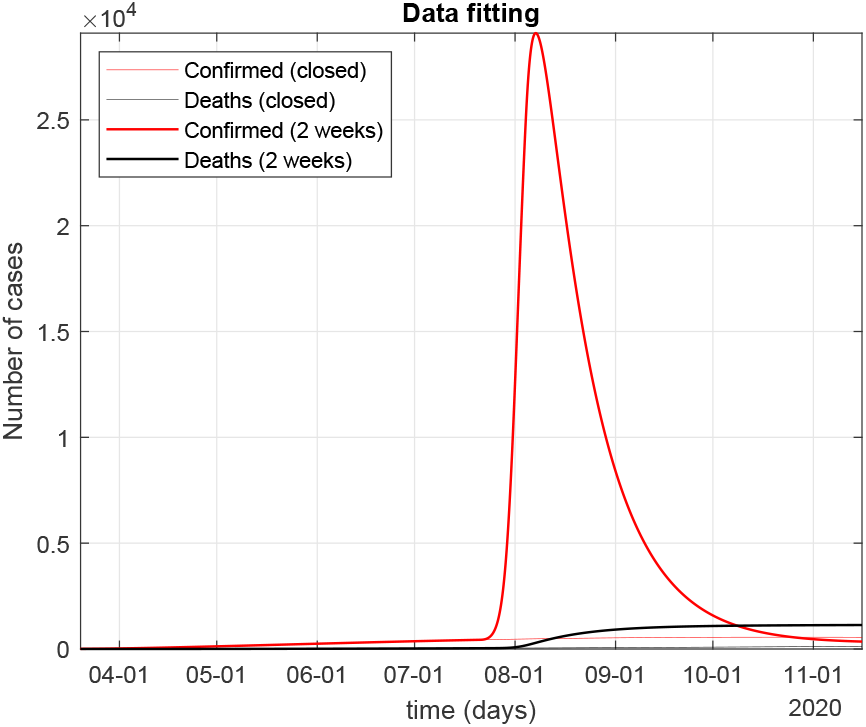
Relax current social distancing 2 weeks after peak

**Figure 3.6:**
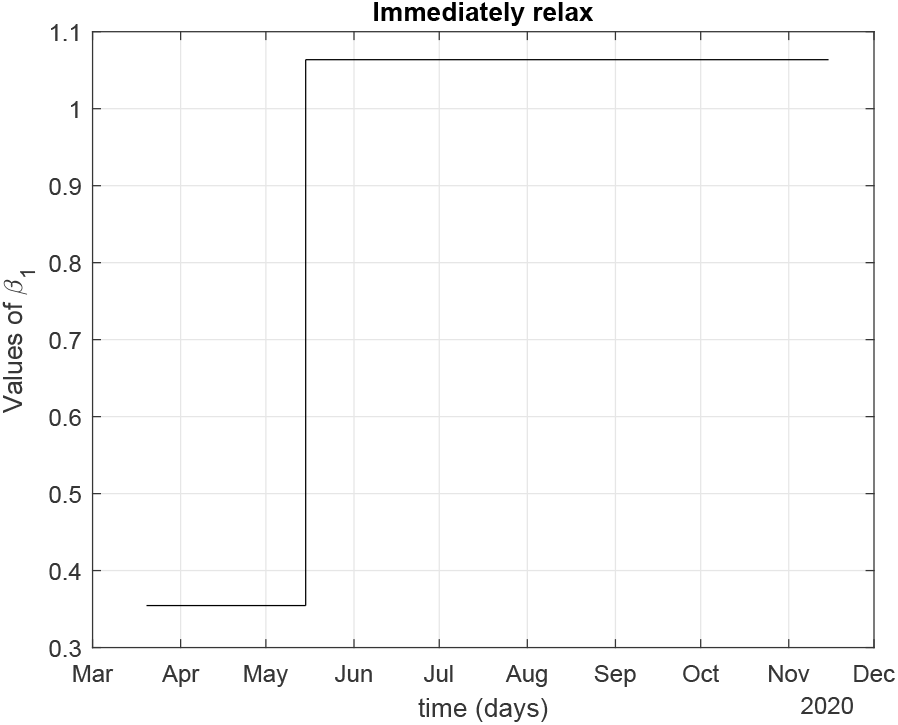
The values of *β*_1_ before and after relax

**Figure 3.7:**
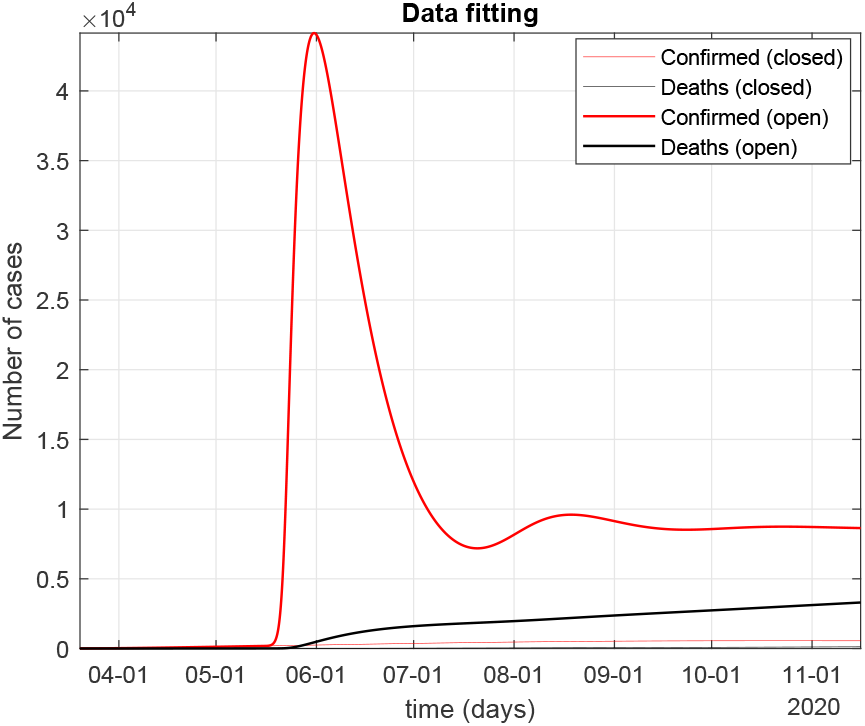
Immediately relax all current restrictions on workplaces

### Remark 3.1.

*In the case of school and workplace closures, we assume that the parameters in the model will remain the same until May 15, i.e*., *β*_1_ = 0.3546, *β*_2_ = 0.6083. *Suppose the social distance is relaxed, the probability of infection becomes doubled. The probability of infection becomes tripled if all restrictions are removed*.

### Remark 3.2.

*Since some people can be treated in isolation at home after infection, we assume a hospitalization rate of 20% of those quarantined cases*.

### Remark 3.3.

*According to our understanding, we assume the peak is the maximum number of new cases per day. According to the prediction curve, the maximum number of new cases per day is 19 on July 21*.

### Remark 3.4.

*As shown in Fig. 3.3, the number of new cases was at least 18 per day before November 15, which had not reached 1% of the peak. So the scenario for intervention (3) was the same as for intervention (1) until November 15*.

## 4 Conclusions and Future Works

From the data, we found that the infection rate in the test area is still high, that is, it is far from enough to just close schools and workplaces, only the implementation of strict city closure measures and maintain strict social distance are the key factors to reduce the number of infected cases and deaths, such as Wuhan in China [2]. For the four different intervention scenarios, we can see that the total diagnosed population did not have a particularly large jump in the case of a continued close, and ended up staying at 3,231. In the two weeks after the peak and in the open scenario, there is a huge jump in the number of infections, which also puts pressure on medical resources.

In future work, we can combine more data to further calibrate our model parameters so that the model can more accurately describe the spread of infectious diseases and predict future trends.

## Data Availability

It is given in the paper (URL) so the results of this report are reproducible.

http://midasnetwork.us/mmods/

